# Improved COVID-19 Serology Test Performance by Integrating Multiple Lateral Flow Assays using Machine Learning

**DOI:** 10.1101/2020.07.15.20154773

**Authors:** Cody T. Mowery, Alexander Marson, Yun S. Song, Chun Jimmie Ye

**Affiliations:** Medical Scientist Training Program, University of California, San Francisco, CA 94143, USA; Biomedical Sciences Graduate Program, University of California, San Francisco, CA 94143, USA; Department of Microbiology and Immunology, University of California, San Francisco, CA 94143, USA; J. David Gladstone Institutes, San Francisco, CA 94158, USA; Innovative Genomics Institute, University of California, Berkeley, CA 94720, USA; Diabetes Center, University of California, San Francisco, San Francisco, CA 94143, USA; Department of Medicine, University of California, San Francisco, San Francisco, CA 94143, USA; Helen Diller Family Comprehensive Cancer Center, University of California, San Francisco, San Francisco, CA 94158, USA; Parker Institute for Cancer Immunotherapy, San Francisco, CA, USA; Chan Zuckerberg Biohub, San Francisco, CA, USA; Institute for Human Genetics, University of California, San Francisco, San Francisco, CA, USA; Computer Science Division, University of California, Berkeley, Berkeley, CA 94720, USA; Department of Statistics, University of California, Berkeley, CA 94720, USA; Division of Rheumatology, Department of Medicine, University of California, San Francisco, San Francisco, CA, USA; Institute of Computational Health Sciences, University of California, San Francisco, San Francisco, CA, USA; Department of Epidemiology and Biostatistics, University of California, San Francisco, San Francisco, CA, USA

## Abstract

Mitigating transmission of SARS-CoV-2 has been complicated by the inaccessibility and, in some cases, inadequacy of testing options to detect present or past infection. Immunochromatographic lateral flow assays (LFAs) are a cheap and scalable modality for tracking viral transmission by testing for serological immunity, though systematic evaluations have revealed the low performance of some SARS-CoV-2 LFAs. Here, we re-analyzed existing data to present a proof-of-principle machine learning framework that may be used to inform the pairing of LFAs to achieve superior classification performance while enabling tunable False Positive Rates optimized for the estimated seroprevalence of the population being tested.

## Introduction

The SARS Coronavirus-2 (SARS-CoV-2) has emerged rapidly and precipitated the Coronavirus Disease 2019 (COVID-19) pandemic that continues to threaten vulnerable populations and disrupt daily life [5]. Although definitive evidence of antibody-mediated protective immunity against SARS-CoV-2 infection is still needed [10, 14], promising early results from trials of convalescent plasma therapy [4] and animal re-infection models [2] raise hopes that antibodies can confer some degree of protection. Because infected individuals nearly uniformly mount detectable serological responses against SARS-CoV-2 [9], sensitive and specific measurement of anti-SARS-CoV-2 serostatus is critical for obtaining accurate estimates of natural immunity (prevalence), as well as infection rates (incidence). Thus, reliable serology tests may provide important epidemiological information to model viral spread and inform non-pharmaceutical interventions including physical distancing and contact tracing.

A number of immunochromatographic lateral flow assays (LFAs) and enzyme-linked immunosorbence assays (ELISAs) were developed swiftly to detect antibodies against SARS-CoV-2 antigens. Recent work by our group and others has revealed potentially inadequate sensitivity and specificity of some of these LFAs [1, 6, 16], suggesting that uninformed usage of these tests could result in inaccurate estimates of seroprevalence or release of misleading information to tested individuals. Although select LFAs perform relatively well, no single LFA is both perfectly sensitive and specific. ELISAs tend to perform better, but they require specialized laboratory equipment that limit their widespread adoption. Because LFAs remain accessible and can be deployed in point-of-care settings, rational LFA deployment may improve diagnostic performance while retaining scalability and ease of use.

Clinical testing methods incorporating multiple laboratory assays achieve superior performance by leveraging the unique strengths of different assays, as is standard practice for HIV testing [8]. Because LFAs utilize a range of antigens and chemistries, we hypothesize that testing with pairs of SARS-CoV-2 LFAs may classify specimen serostatus better than single LFAs. To test our hypothesis, we compare the performance of single LFAs with that of LFA pairs using a simple strategy requiring positive results from both LFAs (*AND* logic). Although the *AND* logic strategy is able to reduce the false positive rate (FPR), it is accompanied by a substantial reduction in true positive rate (TPR) (i.e., sensitivity or power), in some cases to levels below the performance of individual LFAs.

To overcome the limitations of the simple *AND* logic strategy, we demonstrate a proof-of-concept machine-learning classifier that combines the information of semi-quantitative readouts from both IgM and IgG tests to control the FPR at a targeted level while achieving higher TPRs than individual LFAs. Importantly, our classifier obtained the largest TPR gains when low-performing LFAs are combined, thus significantly expanding their utility. The ability to tune the FPR could enable the deployment of LFA pairs across a range of prior probabilities of seropositivity, and facilitate sound statistical comparisons of different tests. We offer a principled framework that may be used to identify well-performing LFA pairs for studies of individual- and population-level immunity, effectively expanding the SARS-CoV-2 immunity testing options to increase testing scalability and distribute supply demands across multiple vendors.

## Results

We re-analyzed recently generated data [16] that examined the performance of SARS-CoV-2 LFAs from 10 vendors (19 tests based on separate IgM and IgG detection for 9/10 assays) scored by two independent readers (Table S1) using a validated semi-quantitative scoring scale [15] with the positivity threshold of ≥1 (Figure 1a-b: “IgM only” & “IgG only”). Here, we use the term “test” to indicate individual IgM or IgG results, and the terms “LFA” and “vendor” to reference the integrated result of IgM and IgG when interpreted together.

**Figure 1:**
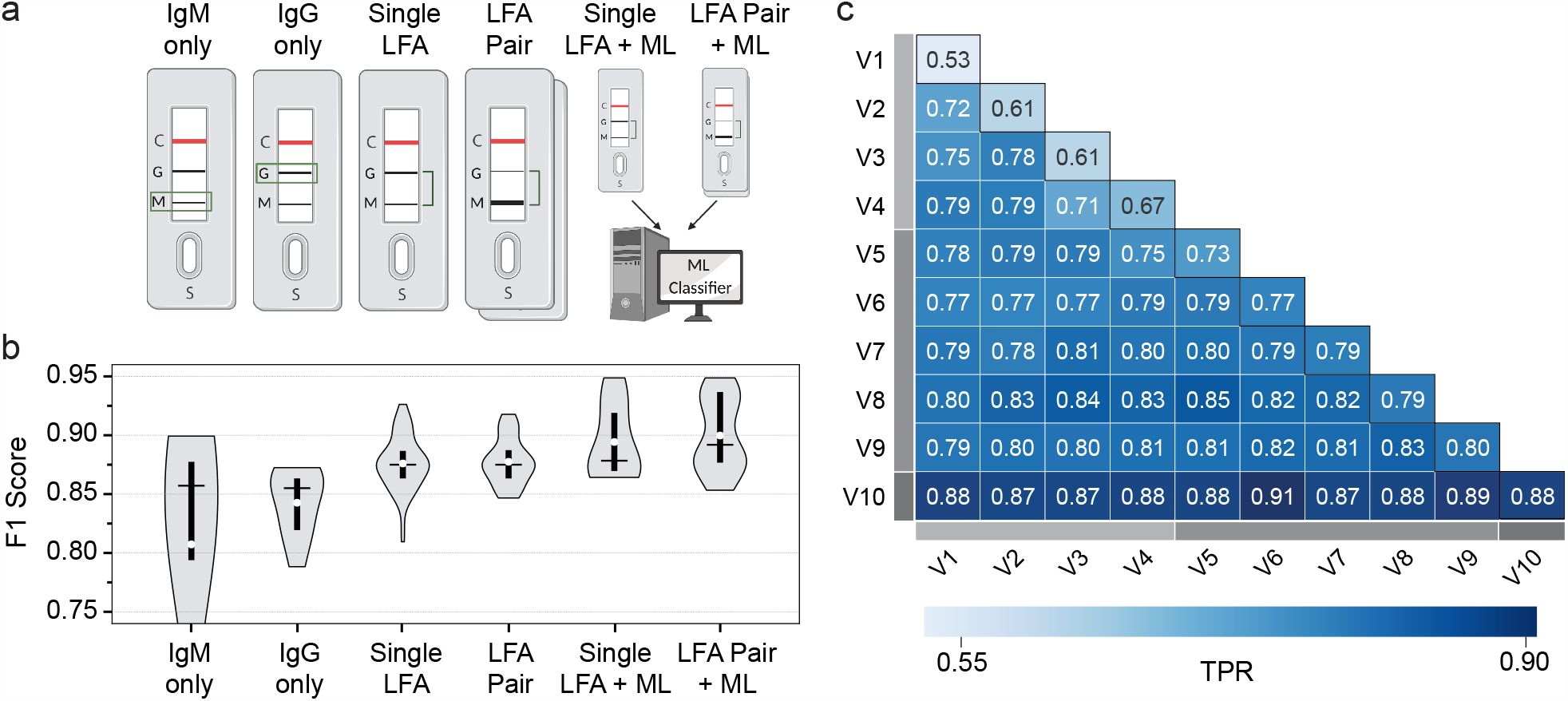
Comparative performance of LFA combination strategies. **a**. Schematic describing data reported in b, including baseline performance characteristics for IgM (“IgM only”, *n* = 9) and IgG (“IgG only”, *n* = 9) tests for each LFA. For single (“Single LFA”, *n* = 10) and paired (“LFA Pair”, *n* = 45) LFAs, specimens were classified as positive if either IgM or IgG test was positive for each LFA. Machine learning (XGBoost) classifier receives both IgM and IgG test information for either single (“Single LFA + ML”, *n* = 10) or paired (“LFA Pair + ML”, *n* = 45) LFAs. **b**. Balanced F-score (or *F*_1_ score) for each experiment outlined in A. One “IgM only” outlier (*F*_1_ = 0.49) is cut off for visualization purposes. The Wondfo LFA was excluded from “IgM only” and “IgG only” because a single band reports signal from both IgM and IgG isotypes. The vertical black bars indicate the range from first to third quartiles, white points indicate mean values, and horizontal bars indicate median values. **c**. Pairwise vendor (V1-V10) TPR performance for XGBoost classifier at *α* = 0.015, binned as low (light grey bar), medium (grey bar), or high (dark grey bar) TPR performance. The diagonal (black outline) specifies TPR results for single LFAs, whereas off-diagonal results reflect TPR of LFA pairs. The reported TPRs were averaged over 100 different random splits of data into 50% training and 50% test sets (see Supplemental Methods).

First, we determined whether a simple LFA pairing strategy improves specimen classification performance as measured by the *F*_1_ score, a well-used evaluation metric in machine learning (see Supplemental Methods). For each LFA, we combined the IgM and IgG results, calling the specimen “positive” if either the IgM or IgG test result is above the positivity threshold (≥1) used in the source publication [16] (Figure 1a-b: “Single LFA”). We find that combining IgM and IgG results for each LFA improves the *F*_1_ score relative to IgM or IgG alone (mean: 88% vs. 81% & 84%, respectively, Figure 1b) by primarily improving TPR (mean: 84% vs. 76% & 77%, respectively, Figure S1). Subsequently, we examined all possible LFA pairs to determine whether requiring concordant positivity of two LFAs (*AND* logic) would improve sample classification. This strategy resulted in no improvements in *F*_1_ scores compared to single LFAs (mean: 88%, Figure 1b) but lowered the TPR (mean: 80%, Figure S1).

This decrease in TPR revealed a vulnerability of the *AND* logic pairing approach to unforeseen negative combination effects, thus motivating us to explore more sophisticated pairing strategies. We evaluated several machine learning classifiers (including random forest, logistic regression, and gradient boosting) using semi-quantitative readouts of LFA test intensities rather than binarized data. We found gradient boosted decision trees (implemented in XGBoost [3], see Supplemental Methods) worked particularly well, so we focused on this approach. Our XGBoost classifier integrates the IgM and IgG test results for each LFA and outputs a probability of positivity for each specimen. Thus, the trade-off between TPR and FPR can be tuned by applying a different probability threshold in accordance with the needs of the user and the prior probability of seropositivity in the test population [16]. Whereas the heterogeneous FPRs reported across different single and *AND* logic-paired LFAs complicates TPR comparisons (Figure S1), controlling the FPR at a desired significance level *α* using a machine-learning classifier enables direct TPR comparisons and, thus, identification of high-performing single and paired LFAs.

We first assessed the overall performance (*F*_1_ score) of the XGBoost classifier at a fixed probability threshold of 0.5. We find that processing single LFAs with XGBoost (mean: 89%, Figure 1a-b: “Single LFA + ML”) outperforms simple single IgM or IgG tests, single LFAs, and *AND* logic LFA pairs mentioned previously. Further, combining LFAs with XGBoost further improves *F*_1_ scores (mean: 90%, Figure 1a-b: “LFA Pair + ML”). Leveraging the aforementioned ability to tune the FPR, we next examined the TPR performance for individual LFAs at fixed significance levels *α* = 1.5% (Figure 1c: diagonal, see Supplemental Methods), 3% (Figure S2a), and 4.5% (Figure S2b). At *α* = 1.5%, we found that XGBoost roughly segregates LFAs from different vendors into three TPR ranges: low (< 70%, light grey bar), mid (70–80%, grey), and high (> 80%, dark grey). Pairing different LFAs with XGBoost (mean: 81%, Figure 1c: off-diagonal) achieves higher TPRs than single vendor XGBoost classifiers (mean: 71%, Figure 1c: diagonal) at the same FPR threshold. We found that vendors that perform well individually (e.g., Vendors 7 & 8) perform marginally better in combination (82% combined vs. 79% & 79% individually). Importantly, LFAs that are lower performers alone (e.g., Vendors 2 & 3) can be paired to achieve significant performance gains over each individual LFA (78% combined vs. 61% & 61% individually) and/or confer modest gains on already mid-performing LFAs (e.g., Vendor 8: 79% individually vs. 83% with Vendor 2 and 84% with Vendor 3). Similarly, two mid-performing LFAs (e.g., Vendors 5 & 8) could be paired to achieve performance in the range of single high-performing LFAs (85% vs. 73% & 79% individually). These effects are not merely additive. For example, certain LFAs enhance the performance of Vendor 10 more than others despite mid-level performance alone; e.g., at significance level *α* = 1.5%, Vendor 9 (80%) performs better than Vendor 6 (77%) individually, but combining Vendors 6 and 10 (91%) is better than Vendors 9 and 10 (89%). These results demonstrate a proof-of-concept implementation of a machine-learning classifier that can effectively identify specific LFA pairs with better classification performance overall and increased sensitivity at a tuned False Positive Rate.

## Discussion

Here, we have demonstrated the utility of machine learning to enhance performance and inform deployment of lateral flow assays (LFAs) for anti-SARS-CoV-2 antibodies. LFAs will likely be integral for accurate estimation of population seroprevalence to inform public health directives, especially in settings where specialized equipment is unavailable [13]. We found that training an optimized gradient boosted decision tree algorithm on LFA LFA Pairs has higher classification performance (*F*_1_ score) than single LFA tests and a more naive LFA pairing strategy. Though LFAs for anti-SARS-CoV-2 antibody detection are likely to improve with time, our framework provides an alternative LFA deployment strategy until a “perfect” SARS-CoV-2 immunoassay is widely available. This computational approach will likely improve the performance of other immunoassays, including SARS-CoV-2 rapid antigen tests and serological assays for other conditions, though the method should be thoroughly validated on a case-by-case basis.

In addition to its superior performance, one of the primary advantages of using a machine learning classifier is the ability to tune the target False Positive Rates in accordance to the population in which the LFAs are being deployed. Given the geographic variability of SARS-CoV-2 prevalence [5], a more stringent FPR may be implemented in low prevalence settings where the pre-test probability is exceedingly low. Conversely, high prevalence populations may be more effectively screened by implementing a classifier that prioritizes higher TPR at the cost of specificity. Such threshold tuning is dependent upon the use of a (semi)quantitative LFA scoring strategy [16], as categorical input data (e.g., “Positive” or “Negative”) cannot be effectively optimized by the classifier. Objective LFA scoring in the form of automated densitometry or an image processing algorithm would be ideal to provide continuous scoring data on which a machine learning classifier can be trained, but, in the absence of this technology, we advocate for use of a validated semi-continuous scoring system to be used by trained readers for optimal results.

Our calculations likely underestimate True Positive Rate given the absence of a gold-standard SARS-CoV-2 immunoassay to identify seroconverted patient specimens. As discussed in our previous work [16], the use of SARS-CoV-2 RT-PCR to classify positive and negative specimens (with the exception of historical, pre-SARS-CoV-2 negative samples) almost certainly includes specimens that have not yet seroconverted. Here, we enrich for seropositive specimens by subsetting to specimens collected 10 or more days after symptom onset [6] (see Supplemental Methods), but we do not have sufficient late timepoint data to more stringently select for seropositivity [7, 9].

LFA batch variability, ongoing assay development, and small sample size preclude our ability to nominate specific LFA combinations for real-world implementation. Rather, we propose here a conceptual framework by which healthcare systems and governmental organizations performing independent LFA evaluations can improve the performance of SARS-CoV-2 immunoassays using machine learning. We demonstrate the approach using a popular machine learning classifier trained on a rather small data set. Although this small sample size limits our ability to explore FPRs lower than 1.5% (see Supplemental Methods), our results demonstrate increased TPR gains with combination testing as the targeted FPR level decreases (Figure 1c, S2). We anticipate that using a model trained on larger data should lead to improved performance and further aid researchers in selecting high-utility LFAs from a collection of evaluated vendors. Additional assay information, including the SARS-CoV-2 antigen bait and secondary antibody detection reagents used in each cartridge, will likely further improve performance by identifying co-linearity and, thus, more effectively identify useful LFA combinations by de-prioritizing those unlikely to enhance one another.

Informed combination LFA testing could help to minimize supply chain limitations by spreading the burden of meeting the world’s SARS-CoV-2 testing demand across multiple manufacturers and LFA vendors. In doing so, our work could effectively expand the number of acceptable SARS-CoV-2 immunoassay testing options, serving as a proof of principle demonstrating the utility of combination LFA testing for more accurate determination of anti-SARS-CoV-2 antibody status.

## Data Availability

Data will be made available upon request.

## Acknowledgements

We thank Caryn Bern and Jeffrey Whitman for their invaluable input regarding this work. C.T.M. is supported by the UCSF ImmunoX Computational Immunology Fellowship and NIH T32GM007618. A.M. holds a Career Award for Medical Scientists from the Burroughs Wellcome Fund, is an investigator at the Chan Zuckerberg Biohub and is a recipient of The Cancer Research Institute (CRI) Lloyd J. Old STAR grant. The research of Y.S.S. was supported in part by an NIH grant R35-GM134922 and the Exascale Computing Project (17-SC-20-SC), a collaborative effort of the U.S. Department of Energy Office of Science and the National Nuclear Security Administration. The information presented here does not necessarily reflect the position or the policy of the Government and no official endorsement should be inferred. A.M. and Y.S.S. are Chan Zuckerberg Biohub Investigators, and C.J.Y. is a Chan Zuckerberg Biohub Intercampus Research Award Investigator.

## Supplement

### Methods

True Positive Rate (TPR) is reported with respect to 79 specimens collected from SARS-CoV-2 RT-PCR-positive patients 10 days or more after patient-reported symptom onset. False Positive Rate (FPR) is estimated against 31 specimens from SARS-CoV-2 RT-PCR-negative patients and 108 specimens from pre-July 2018 historical negative controls.

LFAs were scored using a validated 0-6 LFA scoring strategy [15], and a positivity threshold of ≥ 1 [16] was used for non-machine learning results (Figure 1a-b: “IgM only”, “IgG only”, “Single LFA”, and “LFA Pair”). Missing LFA scores for each vendor (0–15.3% of all specimens, mean: 3.4%, SD: 4.6%) from two independent readers were imputed using a *k*-nearest neighbors algorithm [12], and for each sample the average of the two scores was used for downstream analyses. Pre-processing with imputation and score averaging does not significantly improve baseline TPR (*p* = 0.27, Mann–Whitney U test) or FPR (*p* = 0.60, Mann–Whitney U test) performance metrics of tests with missing data (Table S1).

We employed balanced F-score (*F*_1_ score), a widely-used measure of classification performance in machine learning, to compare the performance of different experiments at divergent false positive rates. It is defined as

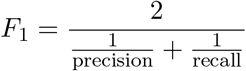

or the harmonic mean of precision (the fraction of true positives among all instances called as positive, or Positive Predictive Value) and recall (TPR, sensitivity, or power).

We implemented an ensemble machine learning classifier using the eXtreme Gradient Boosting (XGBoost) package [3] with ‘`gbtree`’ booster and ‘`binary:logistic`’ objective. This method uses both IgM and IgG test results for each LFA and iteratively generates, evaluates, and refines decision trees to optimize for accurate “positive” or “negative” specimen classification. We trained the XGBoost classifier on 50% of data, used 3-fold cross validation to tune its hyperparameters (`max_depth, min_child_weight, lambda, subsample, colsample_bytree`), and then tested the trained model on the remaining 50% of data. We repeated this experiment 100 times each with different random splits of data into training and test sets, and computed average TPRs at fixed significance levels *α* = 1.5% (Figures 1c), 3%, and 4.5% (Figure S2). Given this train-test split, the lowest possible non-zero FPR that could be considered when testing 50% of the 139 negative specimens is 1*/*(0.5 × 139) ≈ 0.015.

All analyses were performed in Python using the scikit-learn library [11] (except where otherwise specified) and code is available on *Github*.

## Supplementary Table and Figures

**Table S1:** Comparative LFA performance using scores from a single reader (“Reader2”), averaged scores from two independent readers (“Mean”), or averaged scores from two independent readers after imputation (“Imputed”).

|  | False Positive Rate (FPR) |  |  |  |  |  | True Positive Rate (TPR) |  |  |  |  |  |
| --- | --- | --- | --- | --- | --- | --- | --- | --- | --- | --- | --- | --- |
|  | IgM |  |  | IgG |  |  | IgM |  |  | IgG |  |  |
| Vendor | Reader2 | Mean | Imputed | Reader2 | Mean | Imputed | Reader2 | Mean | Imputed | Reader2 | Mean | Imputed |
| Bioperfectus | 0.062 | 0.039 | 0.036 | 0.047 | 0.047 | 0.043 | 0.872 | 0.846 | 0.848 | 0.821 | 0.808 | 0.810 |
| Sure | 0.000 | 0.000 | 0.000 | 0.000 | 0.000 | 0.000 | 0.671 | 0.658 | 0.658 | 0.760 | 0.747 | 0.747 |
| UCP | 0.029 | 0.029 | 0.029 | 0.022 | 0.022 | 0.022 | 0.810 | 0.810 | 0.810 | 0.747 | 0.747 | 0.747 |
| DeepBlue | 0.180 | 0.151 | 0.151 | 0.036 | 0.029 | 0.029 | 0.861 | 0.848 | 0.848 | 0.696 | 0.684 | 0.684 |
| DecomBio | 0.101 | 0.094 | 0.093 | 0.073 | 0.065 | 0.065 | 0.870 | 0.870 | 0.873 | 0.870 | 0.857 | 0.861 |
| Innovita | 0.033 | 0.008 | 0.007 | 0.000 | 0.000 | 0.000 | 0.369 | 0.369 | 0.329 | 0.742 | 0.742 | 0.760 |
| Premier | 0.022 | 0.022 | 0.022 | 0.014 | 0.014 | 0.014 | 0.861 | 0.848 | 0.848 | 0.709 | 0.709 | 0.709 |
| BioMedomics | 0.130 | 0.087 | 0.086 | 0.044 | 0.044 | 0.043 | 0.795 | 0.756 | 0.747 | 0.744 | 0.744 | 0.747 |
| VivaChek | 0.062 | 0.062 | 0.058 | 0.039 | 0.039 | 0.036 | 0.851 | 0.851 | 0.861 | 0.824 | 0.811 | 0.823 |
| Mean | 0.069 | 0.055 | 0.054 | 0.030 | 0.029 | 0.028 | 0.773 | 0.762 | 0.758 | 0.768 | 0.761 | 0.765 |
| SD | 0.058 | 0.049 | 0.049 | 0.024 | 0.022 | 0.021 | 0.155 | 0.152 | 0.165 | 0.055 | 0.051 | 0.053 |
| Vendor | IgM & IgG |  |  |  |  |  | IgM & IgG |  |  |  |  |  |
|  | Reader2 | Mean | Imputed |  |  |  | Reader2 | Mean | Imputed |  |  |  |
| Wondfo | 0.008 | 0.008 | 0.007 |  |  |  | 0.870 | 0.870 | 0.873 |  |  |  |

**Figure S1:**
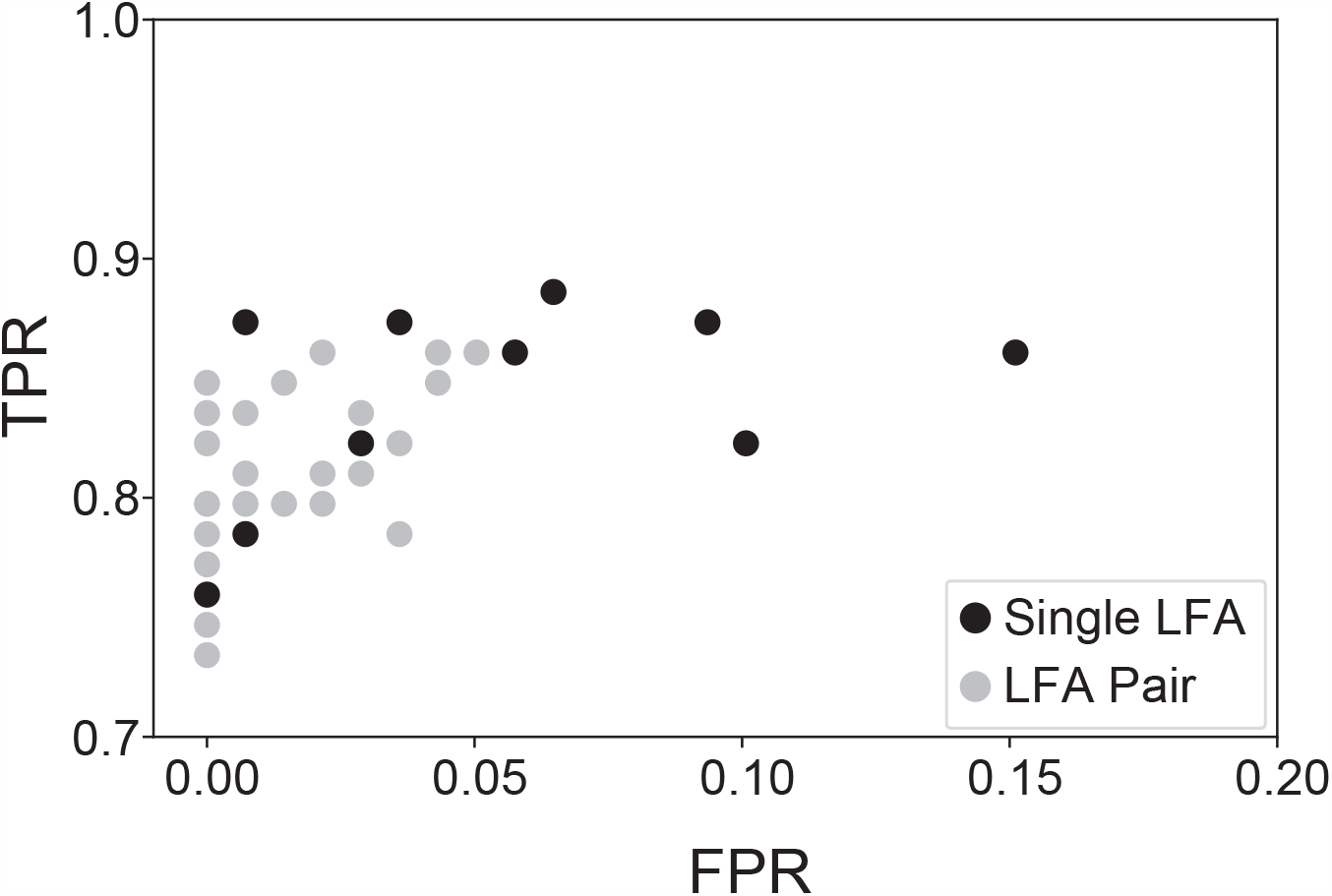
LFA performance with respect to True (TPR) and False Positive Rates (FPR) after integrating IgM and IgG test results for each “Single LFA” (*n* = 10, black). Subsequently, an *AND* logic was applied to require concordant positivity for each “LFA Pair” (*n* = 45, grey) in order to classify a specimen as positive.

**Figure S2:**
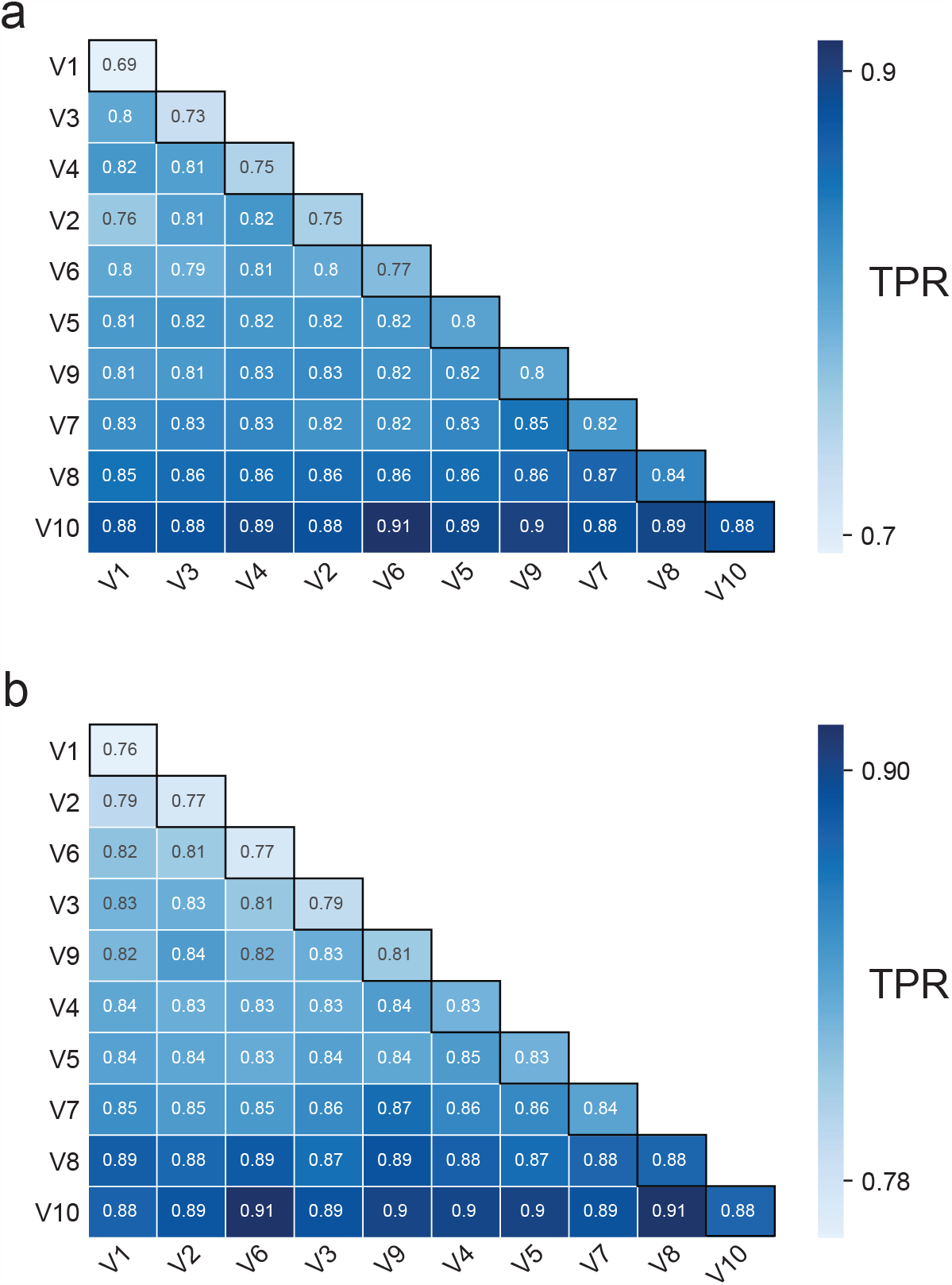
Pairwise LFA performance for XGBoost classifier at significance level *α* = 0.03 (**A**) and *α* = 0.045 (**B**). The diagonal (black outline) specifies results for single LFAs, whereas off-diagonal results reflect TPR of LFA combinations. The reported TPRs were averaged over 100 different random splits of data into 50% training and 50% test sets (see Supplemental Methods).

